# Linking Patient Records at Scale with a Hybrid Approach Combining Contrastive Learning and Deterministic Rules

**DOI:** 10.1101/2025.10.23.25338654

**Authors:** Cheng Cao, Jay Pillai, Sina Ghadermarzi, Sara Daraei

## Abstract

Linking patient records across disparate healthcare systems is essential to create comprehensive views of patient health, yet this task is complicated by inconsistent identifiers, data quality issues, and privacy constraints. Although traditional deterministic and probabilistic methods have been widely used for record linkage, their performance is often limited in the presence of noisy or incomplete personally identifiable information (PII), and privacy-preserving variants commonly restrict matching to exact token equality. This work presents a hybrid record linkage approach, which integrates a deep embedding model with deterministic rules to leverage both the flexibility and noise-robustness of soft embeddings and reliably and predictable baseline performance from deterministic rules. Using a large-scale real-world dataset, a BERT-based embedding model is fine-tuned using a siamese network with contrastive loss to encode PII fields as numeric vectors. This system is implemented and evaluated on a commercial database consisting of 250 million PII records, showing the successful use of the system in a real-world healthcare setting.

## 1 Introduction

As healthcare delivery becomes distributed across a multitude of systems and providers, the need to integrate data from disparate sources has become critical. Many health systems in the United States independently collect and store clinical data about patients, often using locally defined identifiers. As patients interact with multiple healthcare institutions, their information becomes fragmented across these silos, resulting in incomplete views of health records and posing challenges for data-driven healthcare delivery and research [18].

Integrating electronic health records (EHRs) across systems can provide a more comprehensive understanding of individual health and improve clinical decision-making, research, and population health initiatives. Consequently, numerous initiatives have emerged to link health records across systems while preserving data quality and patient privacy [1, 2, 4, 11, 12, 14].

Record linkage (RL), also known as entity resolution, seeks to identify and merge records that refer to the same entity. For matching patients, this process typically relies on personally identifiable information (PII), such as names, birth dates, and ZIP codes. However, PII is both privacy-sensitive

Preprint. and prone to quality issues, including typographical errors and inconsistent or incomplete entries, which complicates direct linkage [8]. As a result recently there has been an intensified interest in privacy-preserving record linkage (PPRL) methods that aim to match records while keeping PII confidential [7]. Common PPRL techniques involve one-way cryptographic hashing of selected PII fields to produce matchable tokens, as employed in networks such as PCORnet [12]. While effective in preserving privacy, these techniques are brittle; exact token matching does not accommodate minor input discrepancies, resulting in diminished recall.

Traditional RL methods are broadly classified as *deterministic*, based on exact matching rules, and *probabilistic*, which use similarity scores to estimate the likelihood of a match [9, 19]. Hybrid methods have also been developed to combine the strengths of both approaches [15]. Recent works often use bloom filters [5, 7, 10, 13, 17], creating a better balance between privacy protection and linkage accuracy, yet challenges persist when applied to noisy real-world data.

Parallel to these developments, machine learning (ML) techniques have been applied to RL tasks, including early work using neural networks [5, 16, 20]. Most ML-based RL models to date, however, rely on relatively simple architectures and small-scale datasets, limiting their practical effectiveness. Transformer-based architectures such as BERT [6] have shown strong performance across various domains by capturing complex relationships in input data through attention mechanisms. Despite this, their application in record linkage remains limited, likely due to the scarcity of large datasets in this domain.

The present study uses a large-scale real-world patient dataset to fine-tune a BERT-based Siamese network [3] using contrastive loss. Patient records are encoded into continuous vector representations (embedding) in which the similarity of the records is reflected by geometric proximity. This embedding-based approach improves robustness to common PII variations—such as misspellings, inconsistent formatting, name changes, and missing values. To harness the strengths of both learned embeddings and deterministic rules, the proposed framework integrates BERT-derived representations with rule-based linkage logic in a hybrid system. This combination achieves strong precision and recall, providing both resilience to data noise and reliability in well-structured matching scenarios.

## 2 Methods

### 2.1 Problem Formulation

Let *D* = {*r*_1_,*r*_2_, *…, r_n_*} denote a dataset of patient records (Section 2.2.1 describes the fields that comprise a patient record). The task of record linkage is to identify all pairs (*r_i_, r_j_*) such that *r_i_* and *r_j_* refer to the same individual. Formally, this represents a binary classification problem over *n*(*n*− 1)*/*2 possible record pairs. Let *M* denote the set of all true match pairs, and 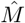 the set of predicted matches. The quality of linkage is evaluated using the following standard metrics:

- Precision: 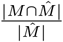
- Recall: 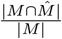
- F1 Score: harmonic mean of precision and recall

#### 2.1.1 Blocking

Due to the computational infeasibility of exhaustively comparing all possible record pairs during linkage, a commonly used strategy of blocking is employed. This strategy partitions the dataset into disjoint subsets *B*_1_, *B*_2_, *… , B_k_*, referred to as *blocks*, based on the value of a blocking token (a specific concatenation of PII fields). The underlying assumption is that when the value of blocking token is different between two records, the chance of real match is very small. Therefore, the computational and statistical benefits of this exclusion outweigh the potential loss of recall due to missed cross-block matches. Consequently, cross-block comparisons—mostly clearly non-matches—are excluded from them model training, and matching process. This strategy significantly reduces the number of candidate pairs while breaking the problem into smaller, independent subproblems.

Multiple blocking tokens, constructed from various combinations of PII fields, were evaluated, :

- T1: first_name[0] + last_name + DOB + gender
- T2: first_name + last_name + DOB + zip_code
- T3: first_name + last_name + gender + DOB
- T4: first_name + last_name + gender

T1 uses DOB, gender, last name, and only the first letter of the first name—yielding high recall and robustness to noise in the name field. While its precision is moderate, it is sufficient for blocking, where recall is prioritized. T3 includes the full first name, improving precision but reducing recall due to added noise. T2 adds zip code, which is unstable over time, further lowering recall. T4 relies solely on full name and gender, lacks DOB, and performs worst due to low discriminative power.

Empirical analysis (Table 1), backed the reasoning above, and choice of T1 as the candidate that achieved the most favorable trade-off between recall and precision.

**Table 1:** Matching Performance for Fuzzy ID (pre-Split/Combine) and Baselines.

|  | **FuzzyID** | Exact | T1 | T2 | T3 | T4 | T1-T4 |
| --- | --- | --- | --- | --- | --- | --- | --- |
| Prec. | **0.94** | 0.98 | 0.81 | 0.97 | 0.95 | 0.004 | 0.97 |
| Rec. | **0.93** | 0.65 | 0.95 | 0.69 | 0.82 | 0.85 | 0.69 |
| F1 | **0.935** | 0.782 | 0.874 | 0.86 | 0.880 | 0.008 | 0.806 |

#### 2.1.2 Unbiased Estimation of Precision and Recall

Since our embedding-based linkage method operates within blocks, and the evaluation dataset comprises only a subset of these blocks, the observed linkage performance may be overestimated. This is because the likelihood of missing true cross-block linkages—those that are ignored due to the blocking strategy—increases as the number of blocks grows. In this section, we present a method to infer an unbiased estimate of the linkage performance across the entire dataset, accounting for all blocks. *Precision* indicates the proportion of assigned fuzzy IDs that correspond to true matches, as verified by available ground truth such as SSNs. It can be estimated without bias from all evaluated record pairs in the evaluation data that involve distinct entities (i.e., excluding self-to-self comparisons), as it depends only on observed pairs within blocks and does not require estimating cross-block matches:

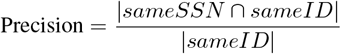

*Recall*, is defined as the proportion of all true matches—regardless of their blocking status—that are successfully linked by the system. Specifically, this includes both the detected matches within blocks and the undetected cross-block matches. The full recall is defined as:

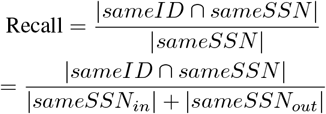

Where:

- *sameSSN_in_*: pairs with the same SSN that reside within the same block.
- *sameSSN_out_*: pairs with the same SSN that are located in different blocks.

Due to the quadratic growth of cross-block same-SSN pairs and the limited sampling of blocks during evaluation, direct recall estimation would be biased. To correct for this, the recall is decomposed into two components:

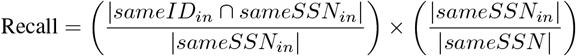

The first term represents within-block recall and can be directly estimated from the evaluation data without bias. The second term reflects the blocking recall, a fixed proportion measurable from the full dataset. This decomposition enables unbiased recall estimation without extrapolation from incomplete block coverage.

### 2.2 Dataset

In this work we use a commercial database that is a combination of records from over 30 healthcare systems across the United States and contain records from nearly a fifth of the US population. The dataset used here is a 20% random sample of this database and thus caputres the diversity of patient information.

#### 2.2.1 Features

Figure 1 presents the 13 available personally identifiable information (PII) fields and their respective fill rates, both before and after data cleaning. Among these, five standard PII fields—first name, last name, date of birth (DOB), gender, and ZIP code—exhibited the highest completeness and were therefore selected as the primary features for the linkage model.

**Figure 1:**
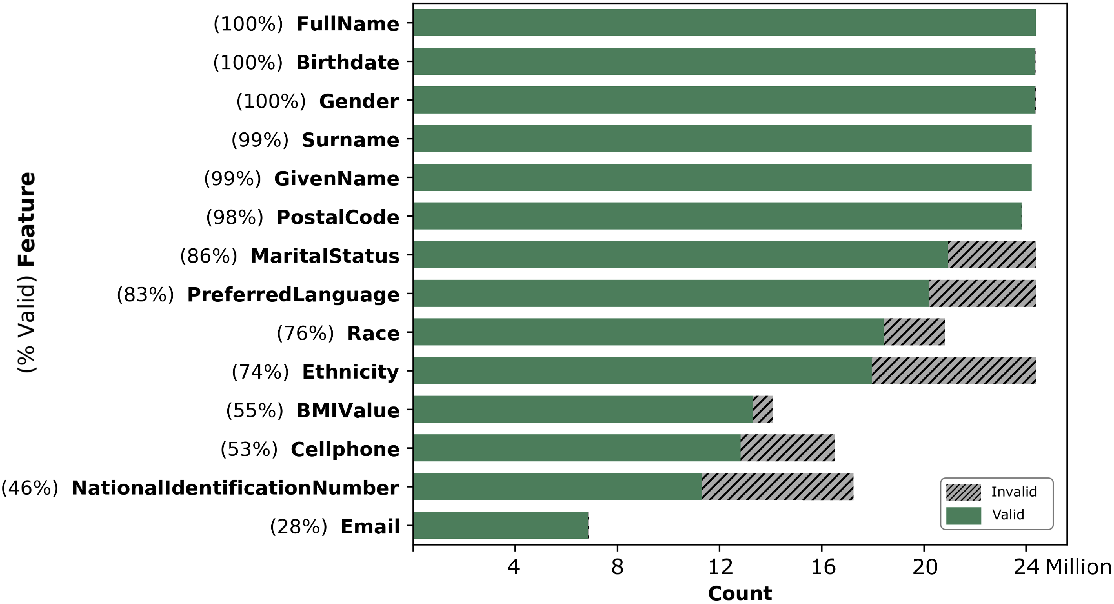
Fill rates for PII features

#### 2.2.2 Ground Truth

Although Social Security Numbers (SSNs) are not universally available and may be affected by data quality issues, valid SSNs—when present—serve as a reliable ground truth anchor. In the absence of manual annotations, valid SSNs are used as a proxy to construct labeled record pairs. To ensure high confidence in the resulting ground truth, after applying a format-check using regular expresions, we manually inspected several hundred SSNs to validate our labeling assumptions. We realized that most of groups of size > 6 are SSNs that are used as a placeholder value (e.g., 111-22-3333) shared by different people. In addition, we observed cases where the same legitimate SSNs were used by multiple members of a family. To filter out these placeholder, or otherwise wrongly shared SSNs, we mark all SSNs that are associated more than one unique DOB and name as invalid. Furthermore, to allow for small variations in DOB and typos in names, we used hamming-distance-based clustering. This process provided a large source of ground-truth labels (1 million records).

#### 2.2.3 Train-Test Split

We assign 20% records to the test set, 10% validation set, and 70% to the training set. To prevent data leakage, the partitioning is performed at the block level, such that all records sharing a blocking token value are assigned to the same partition. This ensures that records in the test set remain disjoint and dissimilar from those in the training set, preserving the integrity of model evaluation.

### 2.3 Embedding Model

A contrastive loss function is employed to train the embedding model:

- Positive pairs (same patient): L2 distance is minimized
- Negative pairs (different patients): L2 distance is maximized (subject to a margin cap)

Training is conducted using a Siamese network architecture, with input pairs sampled within blocks to avoid information leakage. Redundant pairs arising from symmetric permutations are removed, as pair order does not influence the computed distance.

Each patient record is embedded by converting five key PII attributes—first name, last name, date of birth (DOB), gender, and ZIP code—into JSON format, and then passing this structured input into an uncased PubMedBERT model. The model is configured with 24 attention heads, a maximum token length of 1,024, and 16-bit precision. Fine-tuning is performed using a contrastive learning strategy that reduces the Euclidean distance between embeddings for positive pairs (records of the same individual), while increasing the distance for negative pairs (records from different individuals).

The following equation defines the contrastive loss function used:

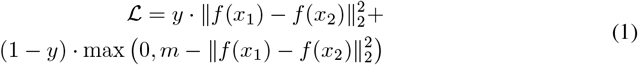

Where:

- *x*_1_,*x*_2_: input pair to the Siamese network.
- *f* (·): embedding function (i.e., network output).
- ∥*f* (*x*_1_) − *f* (*x*_2_)∥_2_: Euclidean distance between embeddings of *x*_1_ and *x*_2_.
- *y*∈{ 0, 1} : label indicating whether the inputs belong to the same class (*y* = 1) or not (*y* = 0).
- *m*: margin parameter enforcing a minimum separation between dissimilar pairs.
- ℒ: computed contrastive loss.

#### 2.3.1 Training

We dedicate 10% of the data to validation for tuning hyper-parameters and we tune them for best AUC on the validation set, with early stopping, where we use the checkpoint from the last epoch where the AUC on the validation set increases (epoch 7).

For contrastive training, we sample positive and negative pairs only within blocks, and only within pairs that don’t have identical PII. Each epoch consist of all possible within-block non-identical pairs, and each batch contain 32 pairs.

### 2.4 Hybrid Linkage Algorithm

Following the embedding of input PII fields, the next step involves clustering the embeddings and assigning a shared identifier to all records within the same cluster. To further enhance linkage accuracy, additional post-hoc correction steps involving high-confidence deterministic rules are introduced, overriding the potential linkage conflicts between existing ids such as valid social security numbers and embedding-based matchings. These steps are implemented within a hybrid framework, as detailed below.

#### 2.4.1 Generate Fuzzy ID (Blocking and Clustering Step)

The generation of fuzzy IDs begins by dividing records into blocks based on a selected blocking token. Within each block, records are clustered based on their embedding similarity, using a defined Euclidean distance threshold. Blocking serves to eliminate low-probability matches early, while also enabling independent and parallel processing within each block.

After clustering, each element in a block is assigned a temporary fuzzy ID, derived by concatenating the block ID (i.e., the blocking token value) with the cluster number. These fuzzy IDs are subsequently refined using overriding rules in the split and combine stages. Algorithm 1 outlines this process, which takes as input a dataset *D* with blocking tokens, embeddings, and a distance threshold.

Clustering is performed by constructing a neighborhood graph where nodes (records) are connected if they lie within the distance threshold. The graph is then partitioned into its largest fully connected subgraphs (cliques), which serve as clusters. This ensures that all pairwise distances within a cluster remain within the defined threshold, thereby avoiding linkage propagation artifacts and ensuring high cluster precision.

##### Algorithm 1

Generate Fuzzy ID

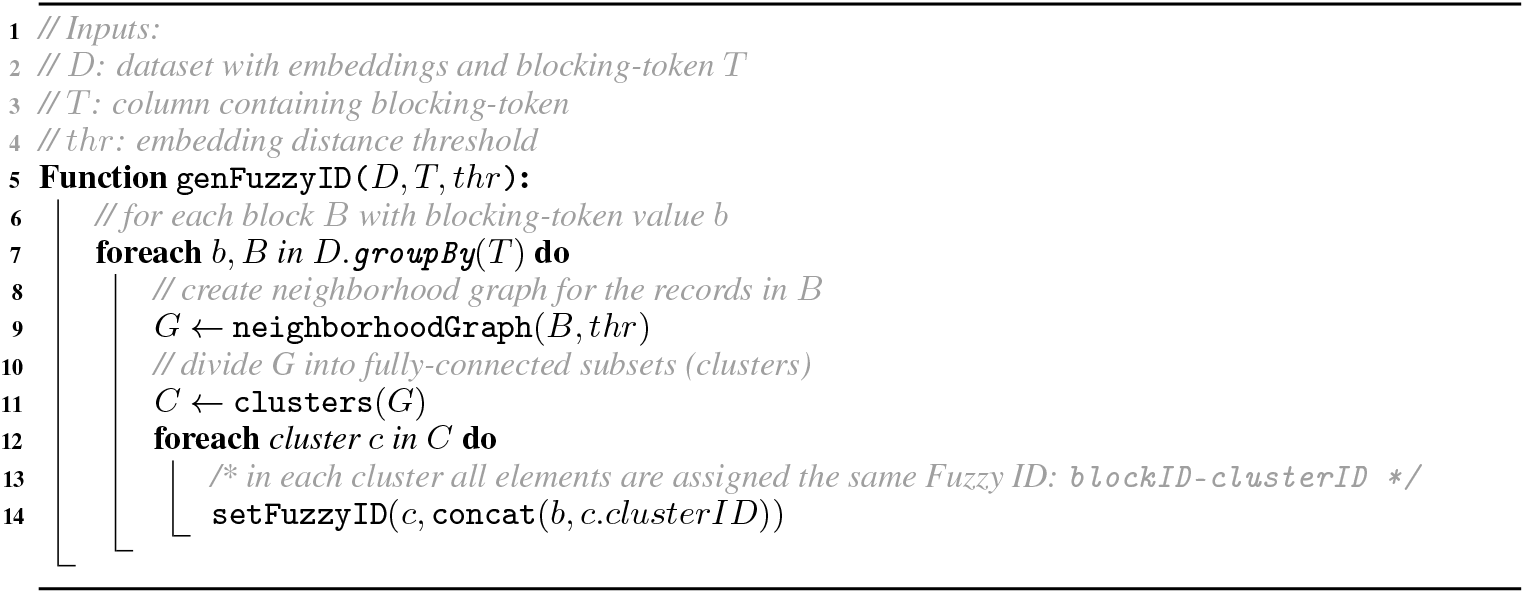

#### 2.4.2 Post-Hoc Corrections Using Deterministic Rules

Once initial fuzzy IDs are assigned, refinement is performed using two types of deterministic rule-based operations:

- Splitting clusters that contain conflicting values for a high-confidence field (e.g., valid SSNs)
- Merging clusters that share a high-confidence field and meet consistency criteria (e.g., same SSN, DOB, and first name)

In the *split* phase, records with different values of a specified *split key* (such as SSN) are required to reside in separate clusters. The presence of differing values for such a key is treated as definitive evidence that the records belong to different individuals. Algorithm 2 describes this procedure, which updates fuzzy IDs based on the values of the split key.

The *combine* phase follows, using a *combine key* (e.g., SSN) for merging. If two records share the same value for a combine key, they are merged into the same cluster. This operation assumes the key provides a sufficient condition for linkage. Algorithm 3 summarizes the process.

Figure 2 illustrates the full hybrid linkage algorithm with an example.

**Figure 2:**
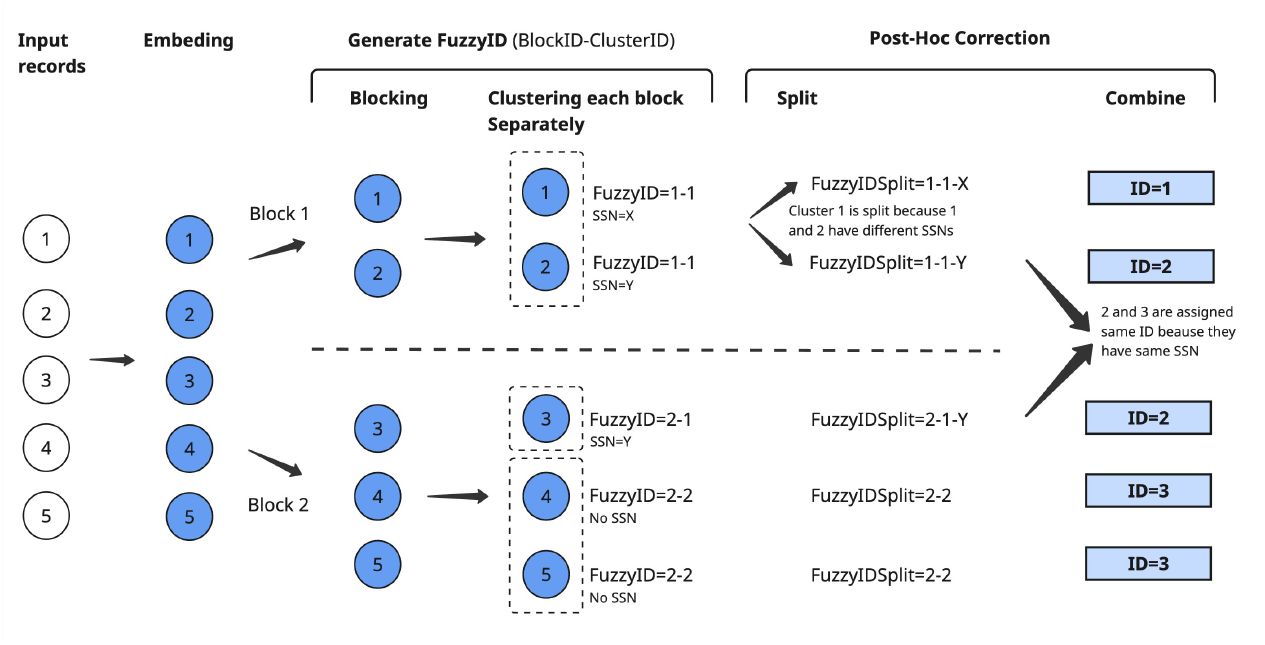
Schematic Illustration of the Hybrid Linkage Algorithm

## 3 Results

### 3.1 Analyzing Embeddings

To evaluate the effectiveness of embedding distances in distinguishing between matching and non-matching record pairs, the distribution of Euclidean distances is analyzed, as shown in Figure 3. The histogram reveals a clear separation between positive and negative pairs, indicating that the learned embedding space successfully encodes semantic similarity. It is important to note that the evaluated pairs belong to the same block and thus already share substantial PII-based similarity.

**Figure 3:**
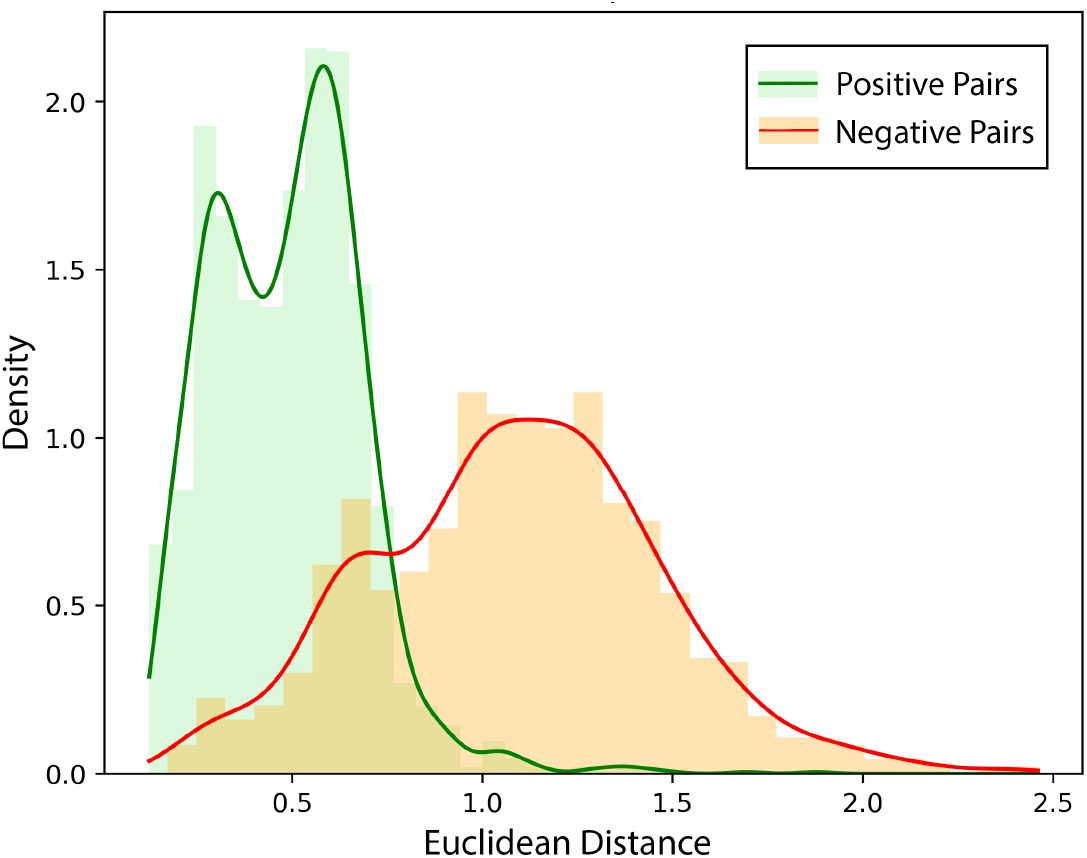
Distribution of embedding distance in positive and negative pairs

#### Algorithm 2

Postprocess-Split

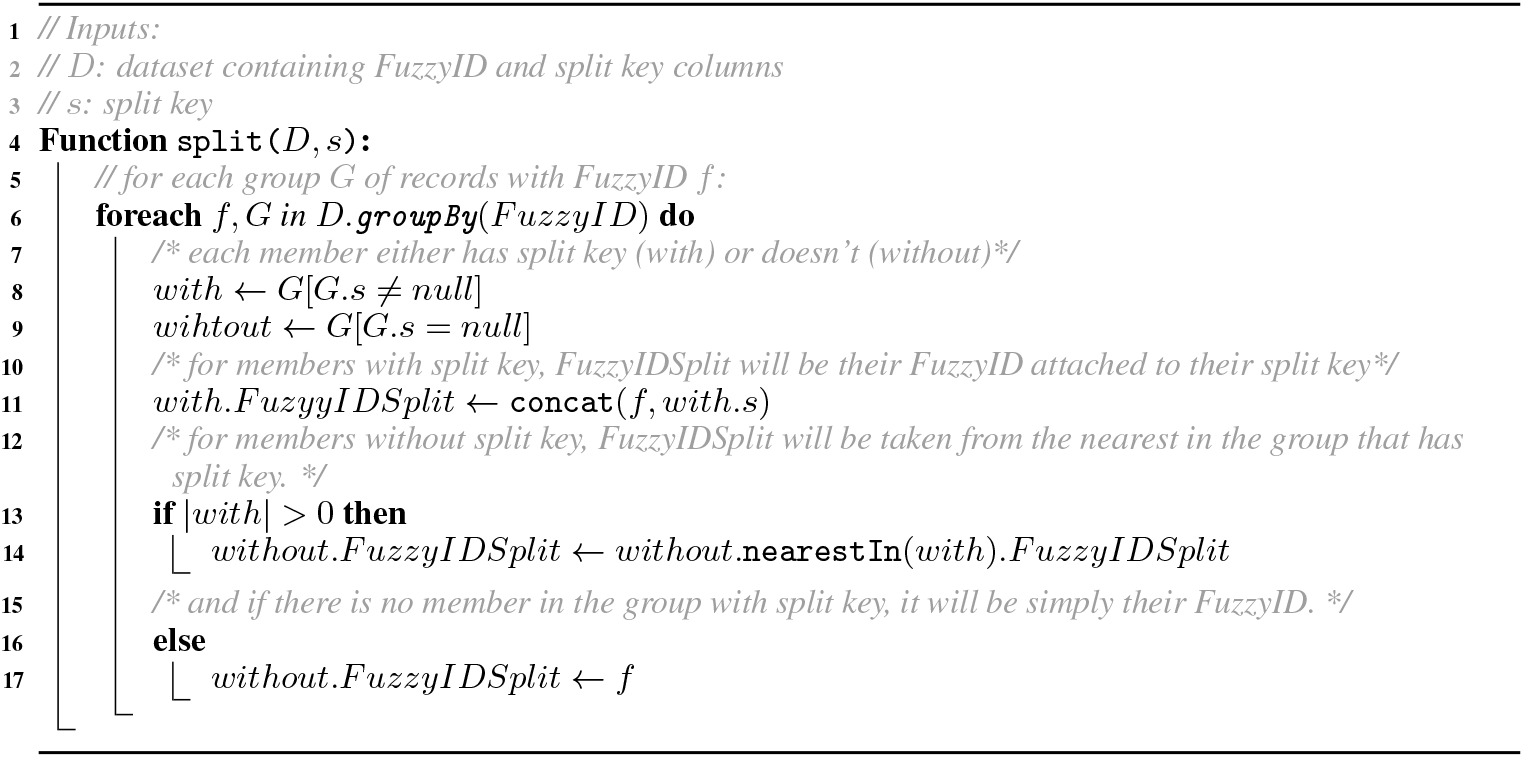

#### Algorithm 3

Postprocess-Combine

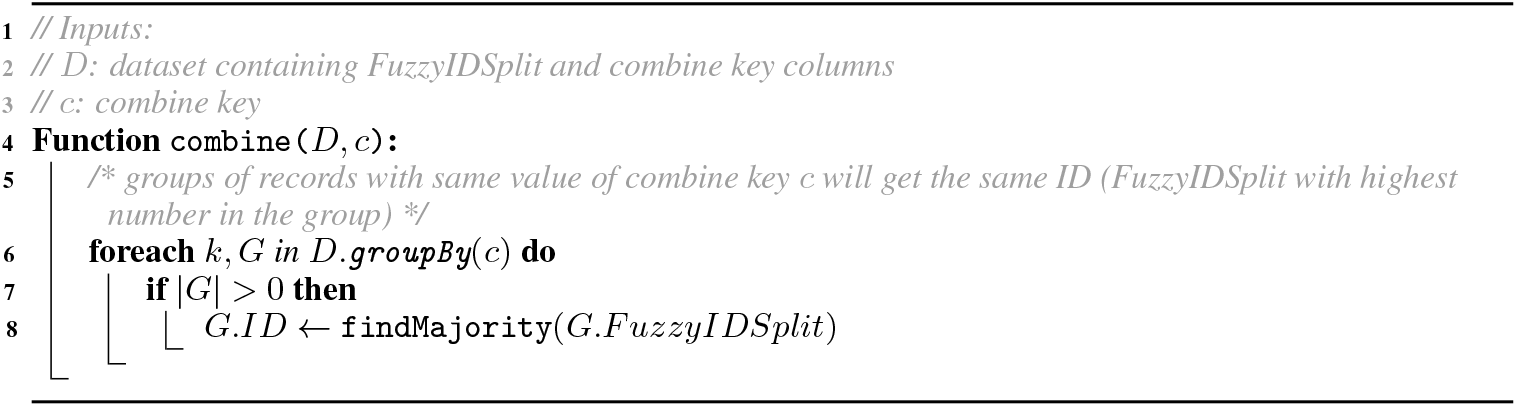

Figure 4 shows an alternative illustration of the discriminative power of embedding distance by plotting the receiver operating characteristic (ROC) curve. The area under the curve reflects strong predictive utility in differentiating matched from unmatched record pairs based on learned distances.

**Figure 4:**
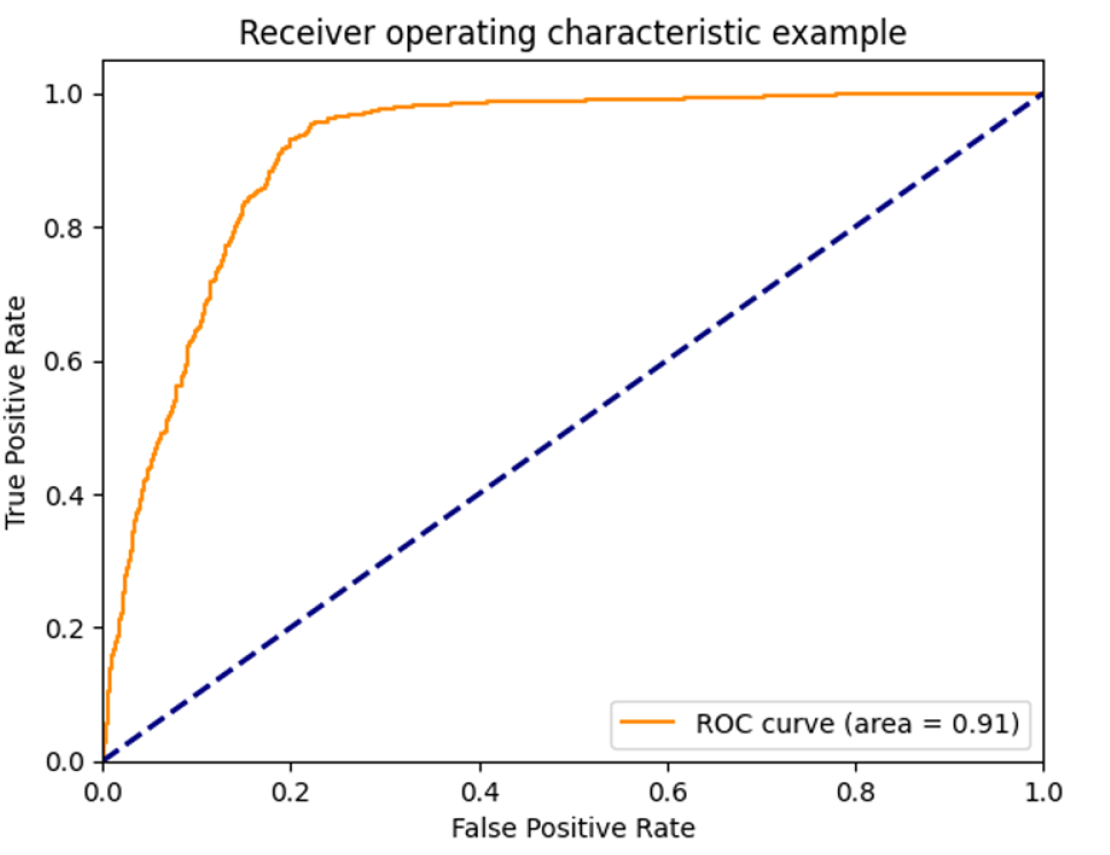
ROC of the embedding distance as a predictor of label

### 3.2 Matching Performance

Reliable evaluation of matching performance typically requires ground-truth annotations, which are often manually curated. Given the scale of the dataset used in this work, manual annotation was infeasible. As an alternative, valid SSNs are employed as a proxy for ground truth during evaluation.

Applying the full hybrid framework—using SSN as both the split and combine key—results in perfect performance, with precision and recall of 100%. However, to isolate the contribution of the embedding-based clustering component, Table 1 reports performance metrics based on fuzzy IDs generated prior to the split and combine stages.

The results demonstrate that the fuzzy ID approach alone significantly outperforms traditional baselines. Precision, recall, and F1 score are reported for various configurations, including exact string match and token-based rules T1–T4 as defined in Section 2.1.1.

*Prec:* Precision, *Rec:* Recall. The column labeled “Exact” reflects exact string match of PII features. The column “T1–T4” represents matching that requires all token rules T1 through T4 to match.

## 4 Discussion

The proposed hybrid framework integrates the adaptability of deep embedding models with the precision and interpretability of deterministic rules. This combination offers a robust solution to challenges associated with data noise, privacy requirements, and computational efficiency in large-scale healthcare record linkage. The learned embeddings can be utilized as privacy-preserving identifiers and may also serve as inputs for secure multiparty computation protocols.

## 5 Conclusion

This study presents a scalable and privacy-aware record linkage framework that leverages contrastive BERT embeddings in conjunction with deterministic rule-based post-processing. The proposed method achieves state-of-the-art accuracy on a large-scale, real-world patient dataset, demonstrating strong generalization performance and robustness in noisy, privacy-sensitive environments.

## Data Availability

Synthetic data can be provided upon reasonable request to the authors

## Notes

### Competing Interest Statement

The authors have declared no competing interest.

### Funding Statement

Project was funded by Truveta Inc.

### Author Declarations

Truveta operates under the HIPAA regulatory framework and for each health system (covered entity), we perform processing under a business associate agreement (BAA) to land, normalize, enhance, transform and de-identify patient records, ensuring that the resulting data meets HIPAA regulatory standards for de-identification and then makes this data available for research.

